# COVID-19 and Acute Kidney Injury in the ICU: Magnitude, Timing, and the Role of Admission Severity in a Large Overlap-Weighted Cohort

**DOI:** 10.1101/2025.08.22.25334252

**Authors:** Anass Ahmed Qasem, Mahmoud Hassanein

**Affiliations:** Department of Internal Medicine and Nephrology, Faculty of Medicine, Zagazig University, Zagazig, Egypt; Prince Sultan Kidney Center, King Salman Armed Forces Hospital, Tabuk, Saudi Arabia; Internal Medicine Department, Faculty of Medicine, Ain Shams University, Cairo, Egypt

## Abstract

**Background:** Acute kidney injury (AKI) is a frequent complication in intensive care unit (ICU) patients and is associated with increased mortality and long-term kidney dysfunction. During the coronavirus disease 2019 (COVID-19) pandemic, high AKI rates have been reported in critically ill patients, but the magnitude and timing of any association in ICU populations remain uncertain—particularly when accounting for baseline severity of illness and missing pre-admission serum creatinine values.

**Methods:** We analyzed 28,612 adult ICU stays (25,923 patients) from the Northwest Intensive Care Unit (NWICU) database between March 2020 and December 2022, excluding patients with end-stage kidney disease (ESKD). COVID-19 status was determined based on laboratory confirmation of SARS-CoV-2 infection when available, or otherwise by ICD-10 diagnosis codes, consistent with clinical practice during the study period. AKI was identified in two post-admission windows—0–48 hours and 0–7 days after ICU admission—using Kidney Disease: Improving Global Outcomes (KDIGO) creatinine-based criteria. Missing covariates were handled using multiple imputation by chained equations (MICE). We estimated the effect of COVID-19 on AKI risk using overlap-weighted logistic regression to balance pre-admission covariates, and repeated analyses adjusting for the Baseline Physiology Index (BPI), a composite severity score derived from the first 12 hours of ICU data.

**Results:** Baseline creatinine was available in 57.7% of ICU stays. In the overlap-weighted cohort (effective sample size ≈ 11,600; maximum absolute standardized difference ≈ 0.01), the standardized 7-day AKI risk was 18.9% (95% confidence interval [CI] 18.5–19.4) in COVID-negative patients and 23.2% (95% CI 22.7–23.7) in COVID-positive patients, corresponding to a risk difference of +4.3 percentage points (pp) (95% CI 3.6–4.9) and an odds ratio (OR) of 1.29 (95% CI 1.24–1.35). Machine-learning-based propensity scores yielded similar results. At 48 hours, no meaningful difference was observed (+0.2 pp; OR 1.02, 95% CI 0.87–1.19). Adjusting for BPI attenuated the 7-day risk difference by approximately 19% in the full cohort and approximately 41% in the BPI-observed subset, but a substantial association persisted.

**Conclusions:** In a large, ICU-admission–anchored cohort balanced on pre-admission traits, COVID-19 was linked to 4–5 extra AKI cases per 100 patients in the first week (not the first 48h). Adjustment for baseline physiologic severity attenuated this risk, indicating partial mediation by illness at presentation, while residual effects suggest other pathogenic pathways. The integrated approach—anchoring at admission, dual-rule KDIGO ascertainment, overlap weighting, and severity-attenuation—offers a robust framework for future COVID–AKI research.

## Introduction

Acute kidney injury (AKI) is a frequent complication in intensive care and is consistently linked to higher mortality and reduced kidney function at discharge [1]. Since the emergence of COVID-19, observational studies have reported substantial AKI rates among hospitalized patients, often with a sizeable fraction reaching KDIGO stage 2–3 or requiring renal replacement therapy [2]. Proposed mechanisms span hemodynamic, inflammatory, microvascular, and treatment-related pathways, but such biologic explanations compete with alternative possibilities: changes in ICU admission patterns, triage delays, or pandemic-era treatment practices that may influence kidney injury risk independently of viral pathophysiology.

For critically ill patients, two uncertainties remain. First, the magnitude of any COVID–AKI association is difficult to estimate because ICU studies often mix heterogeneous populations and admission periods. Second, the timing is unclear: AKI within 48 hours of ICU admission could reflect pre-ICU events or initial resuscitation, whereas AKI emerging later in the first week may arise from ongoing inflammation, evolving multi-organ dysfunction, or treatment exposures.

This timing distinction may be further blurred by diagnostic lag, variable lab schedules, and progression of subclinical injury already present on arrival.

Several methodological challenges complicate inference. A central issue is missing baseline creatinine, which limits KDIGO’s “relative to baseline” definition and risks biased stage distributions if patients without baselines are excluded or if baseline is back-calculated without caution [3,4]. While KDIGO’s absolute criterion (rise ≥0.3 mg/dL within 48 hours) can detect AKI without a baseline [5,6], creatinine-based thresholds in the ICU are susceptible to dilution/concentration effects from fluid shifts and omission of urine output data. A second challenge is confounding: COVID-positive and COVID-negative patients differ in age, comorbidities, admission context, and calendar month, and unmeasured factors—such as care intensity, nephrotoxin exposure, or center-level practice patterns—may further bias comparisons. Causal inference methods that restrict analysis to the “overlap” population, where patients could plausibly belong to either group, can improve balance and precision [7], but cannot guarantee immunity from residual confounding.

We designed this study to address these gaps in a large ICU cohort from the NWICU dataset (March 2020–December 2022) anchored at ICU admission, applying KDIGO creatinine criteria that incorporate both relative and absolute rules to retain patients without a documented baseline, and using overlap-weighted propensity scores to estimate effects in a clinically plausible comparison group with near-perfect covariate balance. We further separated early (0–48 h) and week-1 (0–7 d) AKI within the same causal framework and conducted an explicit attenuation analysis using a pre-admission severity summary (Baseline Physiology Index, BPI) as a mediation probe. Our objectives were: (1) to quantify the 7-day AKI risk difference between COVID-positive and COVID-negative ICU patients in the overlap population; (2) to determine whether any difference is already present within the first 48 hours after admission; and (3) to assess how much of the association is explained by baseline severity. We prespecified that any genuine COVID effect would most likely manifest over the first week rather than immediately, but acknowledge alternative explanations—including system-level factors and treatment pathways—that could produce similar patterns.

## Methods

We performed a retrospective cohort study using the NWICU extracts. Admissions, laboratory, and charted vital-sign tables were merged by the same patient and encounter identifiers (subject_id, stay_id), and timestamps were aligned so that every measurement attached to the correct ICU stay. Time zero was ICU admission. We restricted to adult ICU stays and excluded end-stage kidney disease (ESRD) a priori. After merging and exclusions, the analytic dataset comprised 28,612 ICU stays from 25,923 patients, of which 5.39% were COVID-positive by our primary definition. A patient-specific baseline creatinine was available in 57.7% of stays (median 1.02 mg/dL, IQR 0.78–1.56) and missing in 42.3%; these counts and unweighted AKI rates are summarized in Table 1. We include these numbers in Methods to clarify the size of the analysis set and the extent of missing baseline creatinine, which directly affects how KDIGO rules can be applied.

**Table 1.** Study population and data availability. Among stays with baseline creatinine. †Unweighted cohort rates using KDIGO creatinine rules (relative and absolute criteria).

| Metric | Value |
| --- | --- |
| ICU stays, n | 28,612 |
| Patients, n | 25,923 |
| COVID-positive, % of stays | 5.39 |
| Baseline creatinine available, n / % | 35,701 / 57.7 |
| Baseline creatinine missing, n / % | 26,142 / 42.3 |
| Baseline creatinine (mg/dL), median (IQR)* | 1.02 (0.78–1.56) |
| BPI (Baseline Physiology Index) available, % of stays | ~67 |
| Unweighted AKI by 48 h, % of stays† | 15.4 |
| Unweighted AKI by 7 d, % of stays† | 21.9 |

### Exposure

The exposure was COVID-19 (covid_any), defined as any documented clinical or laboratory evidence of SARS-CoV-2 infection during the hospitalization.

### Outcomes

Acute kidney injury (AKI) was identified using KDIGO creatinine criteria in two windows after ICU admission: 0–48 hours and 0–7 days. For every admission we evaluated (i) a relative-change rule that requires a baseline creatinine (serum creatinine ≥1.5×, 2.0×, or 3.0× baseline for KDIGO Stages 1–3), and (ii) an absolute-change rule that does not require a baseline (increase ≥0.3 mg/dL within any rolling 48-hour window, Stage 1). When baseline creatinine was missing, patients could still be classified as AKI via the absolute rule; the relative rule was only evaluated when a baseline existed. At each timepoint we took the maximum stage reached by either rule and then summarized binary outcomes: any AKI by 48 hours (aki48_any) and any AKI by 7 days (aki7d_any). Unweighted outcome rates appear in Table 1.

### Pre-exposure covariates

To reduce confounding, we used variables determined before or at admission that could influence both COVID status and AKI risk: age, sex, race, admission type, admission location, insurance, comorbidities (chronic kidney disease, diabetes, hypertension, heart failure, chronic liver disease, chronic lung disease, cancer, immunosuppression), and calendar month of admission (numeric year-month) to capture pandemic waves and practice changes.

### Early physiology summary

From the first 0–12 hours after ICU admission we extracted vital signs (heart rate, respiratory rate, mean arterial pressure, temperature, SpO) and labs (sodium, potassium, creatinine, bilirubin, platelets) and built a Baseline Physiology Index (BPI) to summarize “how sick at arrival” (higher = sicker). Each measure was converted to a “derangement” score so that more abnormal values were higher (two-sided variables penalized deviation on both sides), standardized with a robust z-score using the median and median absolute deviation (MAD) (scaled by 1.4826) and capped at ±3 so extremes did not dominate, then averaged across available measures per patient and rescaled to 0–100. Because these 0–12 h values can already be affected by COVID at presentation, we did not include BPI (or those early vitals/labs) in the primary balancing set to avoid adjusting away part of the exposure’s effect; instead, we used BPI later to probe how much of the COVID-AKI link might run through baseline severity (see Table 5 for attenuation analyses and Table 6 for brief definitions).

### Missing data

We created missingness indicators for descriptive reporting (e.g., map_miss, spo2_worst_miss). For analysis, we used multiple imputation by chained equations (MICE) with CART (decision-tree models per variable) to generate m = 5 completed datasets with 10 iterations, excluding identifiers and highly collinear variables from imputation. ESRD exclusions were applied before imputation. Estimates were combined across imputations using Rubin’s rules. Operational definitions (e.g., CART, ESS, ASD) are summarized in Table 6.

### Estimating the effect of COVID on AKI (fair comparisons)

Our target estimand was the effect among patients with overlap—people who could reasonably have been COVID-positive or COVID-negative given their background. First, we estimated each patient’s propensity score (their probability of being COVID-positive) from pre-admission covariates using ridge-penalized logistic regression, allowing flexible (spline) shapes for age and calendar month. Second, we applied overlap weights so that patients who could plausibly be in either group counted more and extremes counted less: weight = PS for COVID− and weight = 1−PS for COVID+. Balance, overlap, and precision diagnostics (absolute standardized differences and effective sample size) are reported in Table 2. Third, on each imputed dataset we fit a weighted logistic regression with AKI ∼ COVID (no other covariates—the weighting already balances them) and used g-computation to obtain standardized risks if everyone were COVID-negative and if everyone were COVID-positive; we then computed the risk difference (COVID+ − COVID−) and the odds ratio, and pooled results across imputations. These primary estimates, alongside a machine-learning propensity sensitivity (XGBoost), are presented in Table 3, with the 48-hour window in Table 4. We also inspected propensity-score distributions by observed COVID status to verify common support before weighting (Figure 1 for the ridge model; Figure 2 for XGBoost). Finally, to examine whether arriving sicker explains part of the association, we re-estimated effects while adjusting for BPI in two ways: in the full cohort using BPI quartiles + a “Missing” category and in the BPI-observed subset using continuous BPI (spline); attenuation of the COVID effect under these specifications is summarized in Table 5.

**Figure 1.**
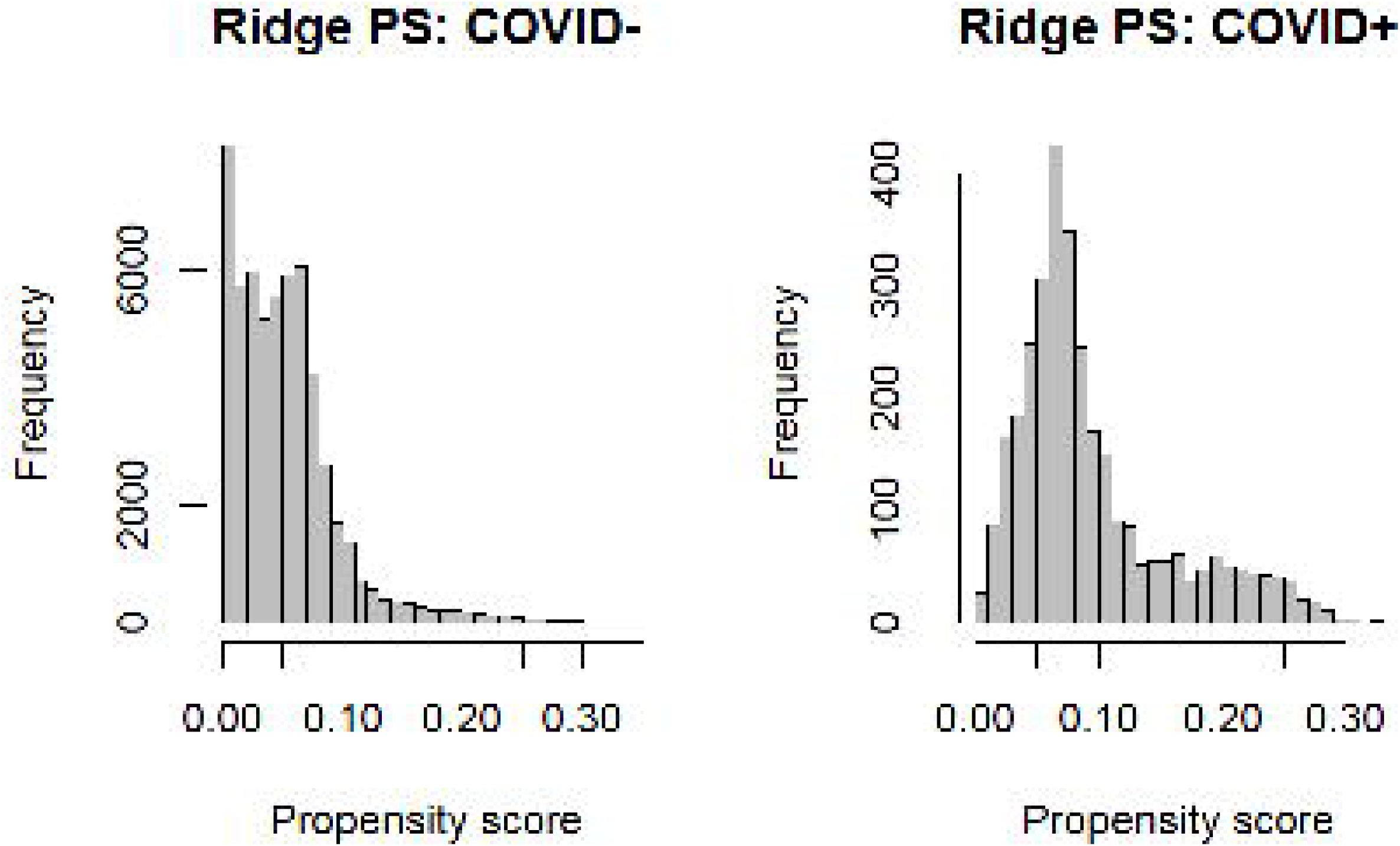
Propensity score distributions by observed COVID status (ridge model). The ridge-model propensity scores (predicted chance of being COVID-positive) are low for most COVID-negative patients and shifted higher for COVID-positive patients, yet the two distributions overlap broadly—roughly in the 0.05–0.20 range—with very few observations above 0.30. Counts are unweighted and the y-axes differ because there are far fewer COVID-positive than COVID-negative stays.

**Figure 2.**
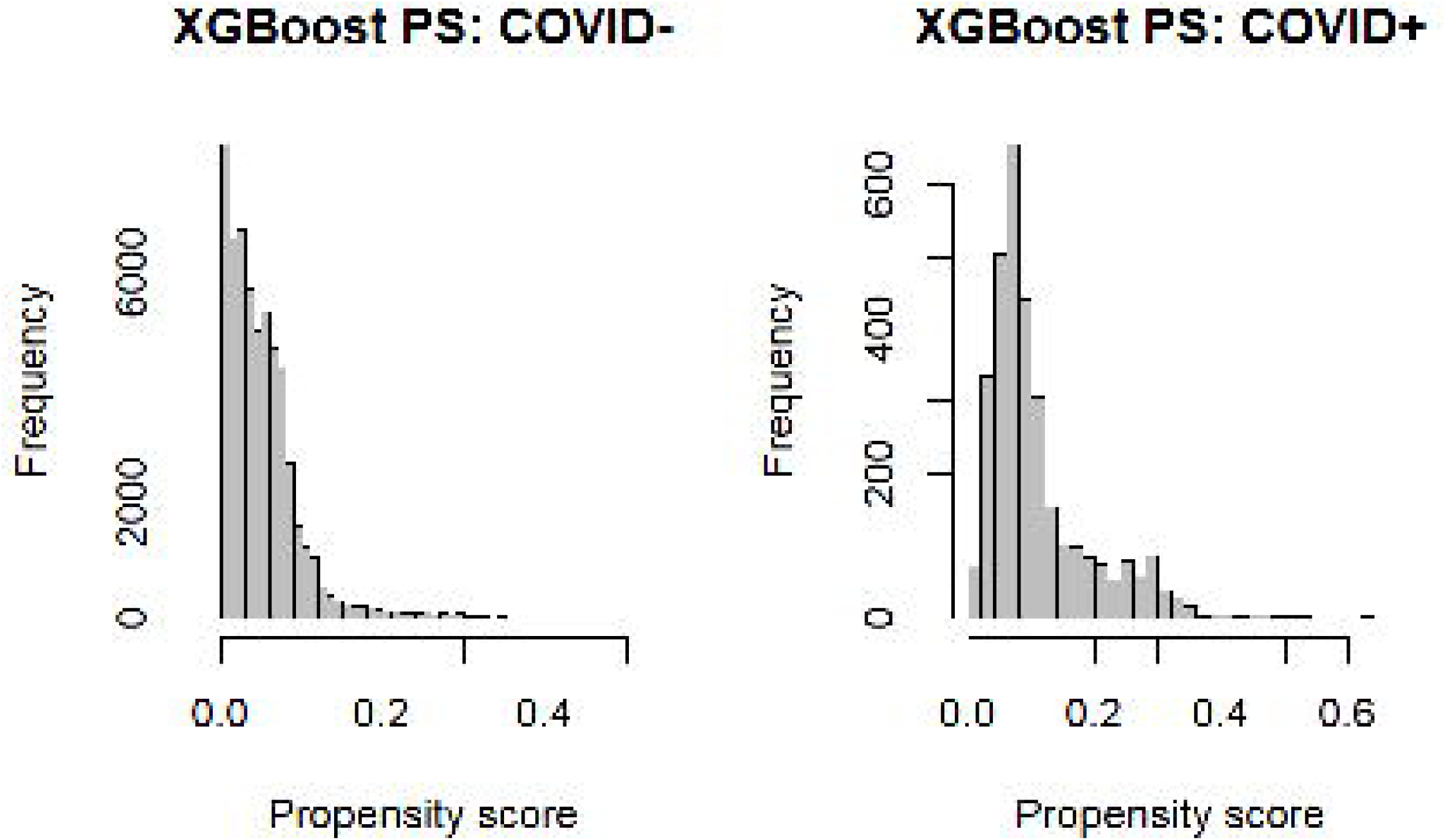
Propensity score distributions by observed COVID status (XGBoost model). Under the XGBoost model, most COVID− patients have low predicted probabilities of being COVID-positive, while COVID+ patients are shifted to higher values; nevertheless, the two PS distributions overlap widely—approximately in the 0.05–0.20 range—with very few extreme scores above 0.30. The counts are unweighted and the y-axes differ because COVID+ stays are much fewer

**Table 2.**
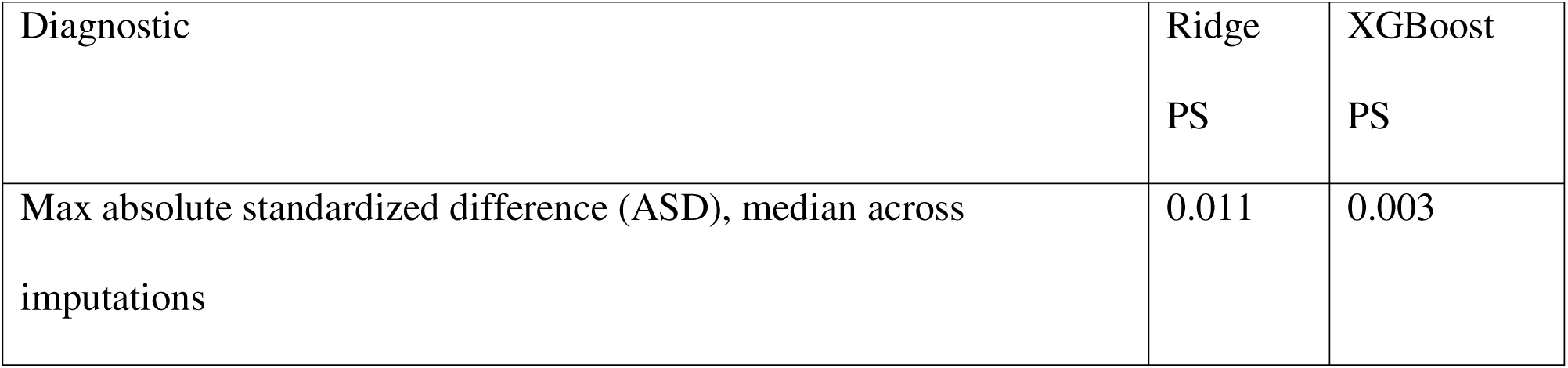
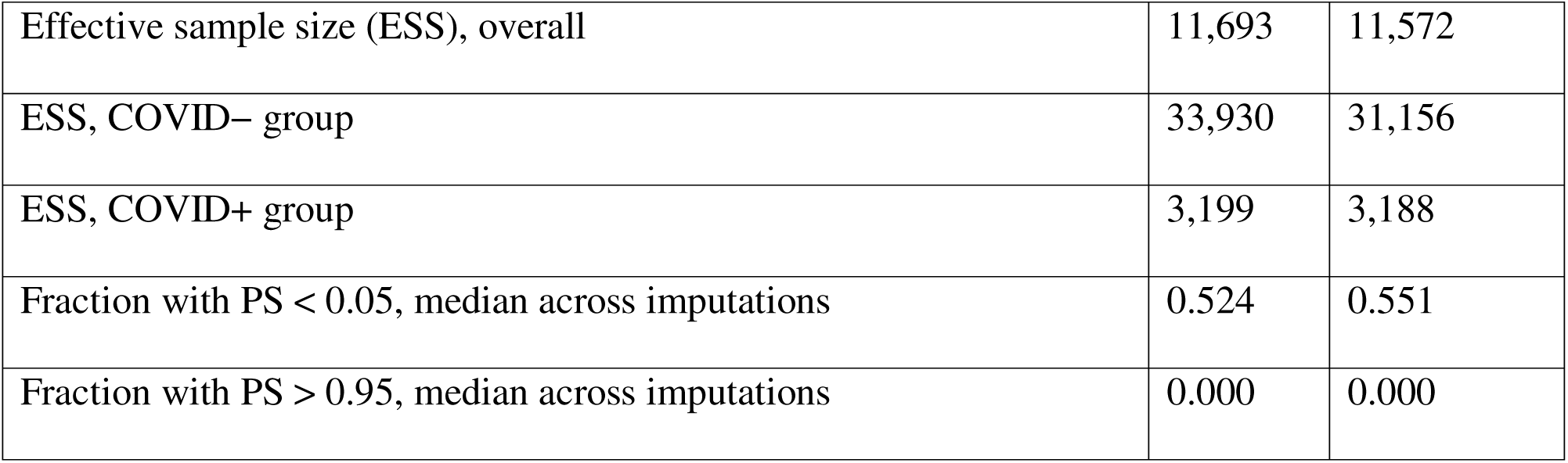
Balance and weighting diagnostics (overlap weighting) ASD < 0.10 indicates negligible imbalance. ESS represents the effective sample size — the number of patients that the weighted data are equivalent to. A larger ESS indicates greater statistical precision

**Table 3.** Primary effect estimates for 7-day AKI (overlap-weighted). pp = percentage points. *E-value computed from the OR; magnitude needed for a single unmeasured confounder to fully explain away the association with both exposure and outcome.

| Estimator | Risk if<br>COVID–, %<br>(95% CI) | Risk if<br>COVID+, %<br>(95% CI) | Risk difference,<br>pp (95% CI) | Odds ratio<br>(95% CI) | E-<br>value* |
| --- | --- | --- | --- | --- | --- |
| Ridge PS →<br>overlap weights | 18.9 (18.5–19.4) | 23.2 (22.7–23.7) | +4.3 (+3.6 to<br>+4.9) | 1.29<br>(1.24–<br>1.35) | 1.90 |
| XGBoost PS →<br>overlap weights | 18.8 (18.3–19.3) | 23.2 (22.7–23.7) | +4.4 (+3.7 to<br>+5.1) | 1.30<br>(1.25–<br>1.36) | 1.93 |

**Table 4.** Early window (48 h) effect estimates (overlap-weighted) No meaningful difference at 48 h; the gap emerges by day 7 (see Table 3).

| Estimator | Risk if<br>COVID–, % | Risk if<br>COVID+, % | Risk difference,<br>pp | Odds ratio<br>(95% CI) |
| --- | --- | --- | --- | --- |
| Ridge PS → overlap weights | 13.0 | 13.2 | +0.2 | 1.02 (0.87–1.19) |

**Table 5.** Severity-adjusted (“direct”) effects using the Baseline Physiology Index (BPI) “BPI quartiles + Missing” = five-level BPI category (Q1–Q4, Missing). “Continuous BPI (spline)” available only when BPI is observed. Different “before” values (4.3 vs 5.0) reflect different analysis sets.

| Analysis set & adjustment | Risk difference, pp<br>(before → after) | Attenuation,<br>% | Odds ratio<br>(before → after) | Attenuation,<br>% |
| --- | --- | --- | --- | --- |
| Full cohort (S2): BPI quartiles + “Missing” | 4.3 → 3.4 | 19 | 1.29 → 1.25 | 13 |
| BPI-observed subset (S1): continuous BPI (spline) | 5.0 → 2.9 | 41 | 1.30 → 1.18 | 38 |

**Table 6.**
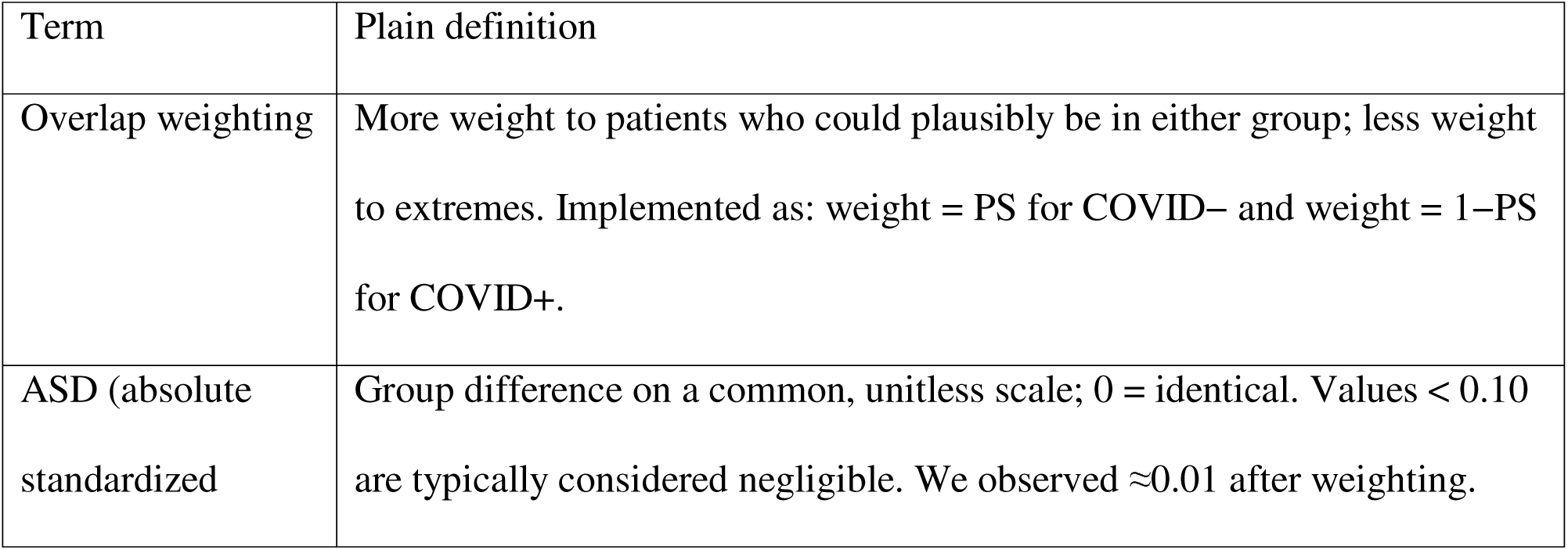
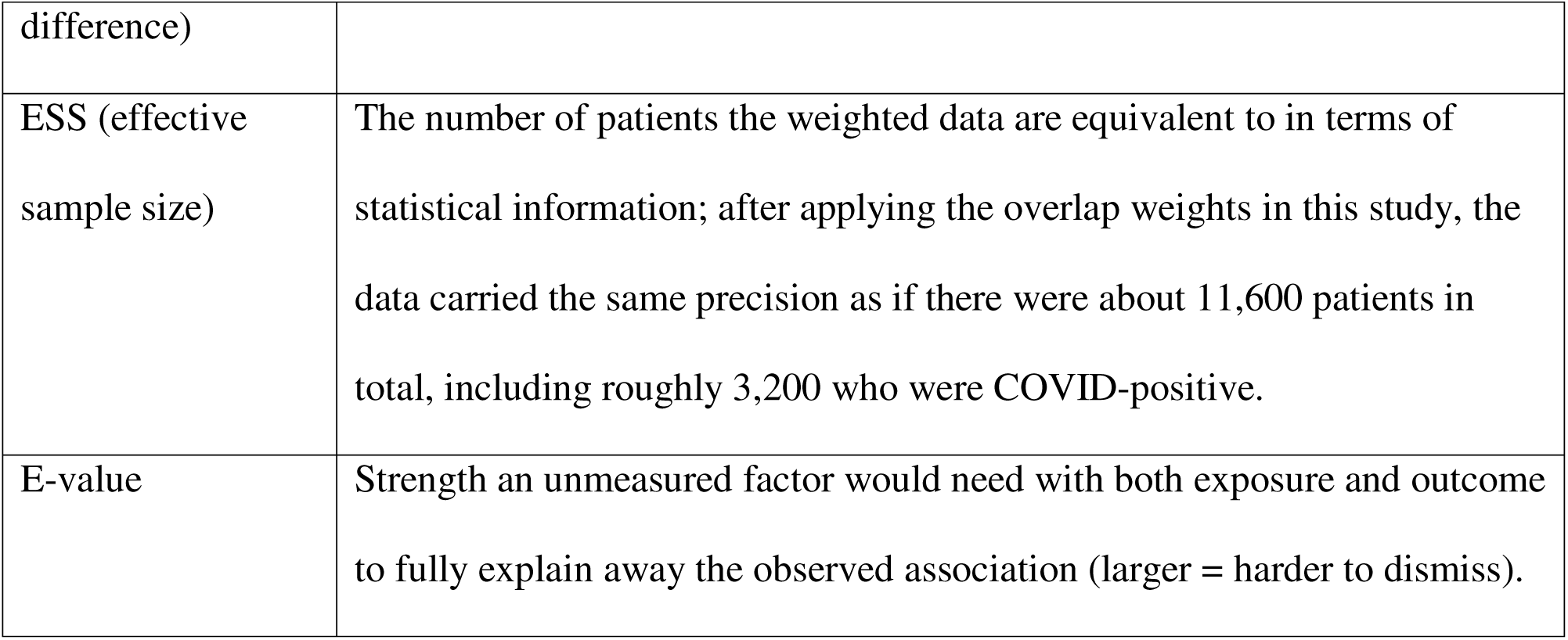
Operational definitions for readers.

### Software and computing environment

Analyses were conducted in R version 4.5.1 (2025-06-13, UCRT build; “Great Square Root”), within the RStudio environment.. Key packages included: mice (imputation), dplyr (data wrangling), zoo/splines (calendar month, spline bases), glmnet (ridge propensity model), xgboost (machine-learning propensity), cobalt (balance diagnostics), and MASS (parametric simulations).

## Results

Patients without a baseline creatinine still qualified for AKI via the KDIGO absolute rule (serum creatinine rise ≥0.3 mg/dL within 48 hours), so missing baseline did not exclude them from detection. In the unweighted cohort, ∼15% developed AKI within 48 hours and ∼22% within 7 days (Table 1).

To enable a fair comparison, we reweighted the data toward patients who could plausibly have been in either group given pre-admission factors (overlap weighting). After weighting, COVID-positive and COVID-negative groups were essentially indistinguishable on those factors: the maximum absolute standardized difference across all covariates was ≈0.011 (median across imputations), well below the 0.10 threshold for negligible imbalance, and the effective sample size remained large (≈11,600 overall; ≈3,200 among COVID-positive), indicating stable estimates drawn from the comparable middle of the cohort (Table 2).

In this balanced overlap population, the standardized 7-day AKI risk was 18.9% (95% CI 18.5– 19.4) if everyone were COVID-negative and 23.2% (95% CI 22.7–23.7) if everyone were COVID-positive—an absolute difference of 4.3 percentage points (95% CI 3.6–4.9) with an odds ratio of 1.29 (95% CI 1.24–1.35). Re-estimating the “chance of COVID” with a machine-learning model (XGBoost) produced essentially the same results (risk difference +4.4 points; odds ratio 1.30), showing the finding does not hinge on how the propensity scores were modeled. The E-value ≈1.93 for the odds ratio indicates that a single unmeasured factor would need ∼1.9-fold associations with both exposure and outcome (after balancing) to fully explain away the association (Table 3). Over 48 hours, there was no meaningful difference (risk difference ∼+0.2 points; odds ratio ∼1.02, confidence interval includes no effect), and the gap emerged by day 7 (Table 4).

We then asked whether patients with COVID simply arrived sicker and whether that explained the association. In the full cohort, we adjusted for baseline physiologic severity by including the Baseline Physiology Index (BPI) as quartiles (Q1–Q4) plus a fifth “Missing” category, retaining patients without BPI. With this adjustment, the 7-day AKI risk difference decreased from 4.3 to 3.4 percentage points (≈19% attenuation) and the odds ratio from 1.29 to 1.25 (≈13% attenuation). In the subset with observed BPI, where we could adjust more finely using continuous BPI (spline), the risk difference fell from 5.0 to 2.9 percentage points (≈41% attenuation) and the odds ratio from 1.30 to 1.18 (≈38% attenuation). The different starting differences—4.3 vs 5.0—reflect the different analysis sets (full cohort vs BPI-observed subset). These severity-adjusted results are summarized in Table 5 (with brief definitions of metrics in Table 6). Taken together, arriving sicker explains a meaningful share of the COVID–AKI difference, but a substantial component remains even after accounting for initial physiologic severity, supporting a genuine increase in 7-day AKI associated with COVID in comparable ICU patients.

## Discussion

In a large ICU cohort balanced on pre-admission characteristics, COVID-19 was associated with roughly 4–5 additional AKI cases per 100 patients by day 7, with no meaningful difference at 48 hours. This pattern suggests processes that develop during the first week of critical illness rather than an immediate admission effect, consistent with prior reports of COVID-related AKI in hospitalized patients [8,9]. The association was robust to how we estimated the propensity scores (ridge vs. machine learning; Table 3) and to stringent balance checks (ASD ≈ 0.01; ESS high; Table 2). Adjusting for baseline physiologic severity explained part of the association (attenuation ∼19% in the full cohort; ∼41% in the BPI-observed subset), but a substantial component remained (Table 5).

The attenuation after adjusting for BPI indicates that some of the COVID–AKI link reflects patients arriving sicker, but the residual effect points toward additional pathways—such as inflammation, hemodynamic instability, microvascular injury, and nephrotoxic exposures— described in consensus statements, pathophysiology reviews, and pathology series [9–11]. The E-value (∼1.9) is consistent with a moderately strong unmeasured confounder being required to fully negate the observed association after accounting for extensive measured covariates and excellent balance [12] (Tables 3 and 6).

We anchored time at ICU admission; used KDIGO criteria that detect AKI even when baseline creatinine is missing via the absolute 0.3 mg/dL in 48 h rule; applied overlap weighting to target a clinically credible population where treated and untreated patients are truly comparable; demonstrated near-perfect covariate balance and high effective sample size; and confirmed stability with a machine-learning PS (Tables 1–4, 6). Visual common support was demonstrated in the PS histograms for both modeling approaches, reinforcing the validity of the overlap estimand (Figure 1; Figure 2). Our two BPI adjustments—quartiles + Missing in the full cohort and continuous BPI in the observed subset—provide consistent evidence that baseline severity explains part but not all of the effect (Table 5).

While elements of our design—such as propensity-based adjustment or stratifying AKI by timing—have appeared in some prior COVID–AKI studies, the present analysis is, to our knowledge, the first ICU-focused cohort to combine (i) an ICU admission time anchor, (ii) KDIGO creatinine criteria incorporating both relative and absolute rules to retain patients without a documented baseline, (iii) overlap-weighted propensity scores with demonstrated near-perfect balance and large effective sample size, (iv) separation of early (0–48 h) and week-1 (0–7 d) AKI within the same causal framework, and (v) an explicit attenuation analysis using a pre-admission severity summary as a mediation probe. This combination addresses common sources of bias—heterogeneous time anchors, baseline creatinine missingness, residual imbalance, and conflation of pre– and post-ICU injury—and yields a methodologically robust benchmark for both historical and future COVID–AKI comparisons.

Baseline creatinine was missing in ∼42%, which influences AKI stage mix (though not detection via the absolute rule) and may correlate with unmeasured factors; we mitigated this by retaining such patients and by using BPI-Missing as its own level (Tables 1 and 5). The overlap estimand emphasizes patients with realistic assignment to either exposure group; results generalize most directly to that overlap population rather than to all ICU patients. A small tail of high or low PS values remained (Figures 1–2), which overlap weighting down-weights but does not eliminate. Residual confounding remains possible despite strong balance metrics; the E-value bounds how strong such a factor would need to be but does not eliminate the risk (Tables 2–3, 6) [12]. Fourth, AKI ascertainment used creatinine-based KDIGO only; urine output was not incorporated and could change incidence estimates. Finally, practice patterns and treatments likely varied across calendar months; we modeled month flexibly in the PS, but residual era effects could remain. In comparable ICU patients, COVID-19 carries a meaningful increase in 7-day AKI risk. Given that the signal is absent at 48 hours and persists after accounting for arrival severity, prevention efforts may need to focus on the first week—e.g., hemodynamic optimization, careful stewardship of nephrotoxins, and early recognition of kidney-injury trajectories in COVID-positive patients [9,11].

Two priorities emerge: (i) time-to-event analyses with competing risks (death, discharge) to map the daily hazard of AKI more precisely; and (ii) treatment-pathway analyses (fluids, vasopressors, antivirals, antibiotics) to identify modifiable drivers of the week-1 AKI signal. Extending BPI to incorporate urine output and adding lab-confirmed COVID as the primary exposure in sensitivity analyses would further refine inference.

## Conclusion

In this large, ICU-admission–anchored cohort, COVID-19 was associated with a modest but clinically meaningful increase in 7-day AKI risk, equivalent to 4–5 additional cases per 100 patients. This excess risk was not evident within the first 48 hours of admission, suggesting processes that unfold during the first week of critical illness. Adjustment for baseline physiologic severity attenuated but did not eliminate the association, implying both illness-at-presentation and other mechanisms contribute. These findings highlight the first week of ICU care as a key window for kidney injury prevention in COVID-19 and establish a robust analytic framework for future investigations into modifiable drivers of AKI in this setting.

## Data Availability

All data produced in the present study are available upon reasonable request to the authors.

https://physionet.org/content/nwicu-northwestern-icu/0.1.0/

## Acknowledgments

The authors used AI-assisted tools during the development of the statistical analysis workflow for this study. Specifically, OpenAI’s ChatGPT was employed to support the generation and refinement of R code within the RStudio environment (R version 4.5.1, “Great Square Root”). The tool assisted with data wrangling, imputation, propensity score modeling, and diagnostics, particularly in relation to packages such as *mice*, *dplyr*, *zoo*, *splines*, *glmnet*, *xgboost*, *cobalt*, and *MASS*. All methodological choices, code verification, and interpretation of results were determined and critically reviewed by the authors.

## Author Contributions

A.A.Q.: Conceptualization; Methodology; Supervision.

M.H.: Data processing; Software; Statistical analysis; Writing – review & editing.

